# Evaluation of School Vaccine Coverage and Medical Vaccine Exemptions Following the Repeal of School Entry Nonmedical Vaccine Exemption Options in New York State

**DOI:** 10.1101/2023.08.17.23294227

**Authors:** John W. Correira, Rhiannon Kamstra, Nanqing Zhu, Margaret K. Doll

## Abstract

**Importance:** Following the repeal of nonmedical vaccine exemption options from school entry immunization laws in California, gains in vaccine coverage were partially offset by increases in medical vaccine exemptions. Although several U.S. states, including New York State (NYS), recently adopted similar legislation, the impact of these laws on school vaccine coverage and medical vaccine exemptions has not yet been examined.

**Objective:** To estimate the effects of NYS legislation eliminating nonmedical school entry vaccine exemptions on required vaccine coverage and the uptake of medical vaccine exemptions at NYS schools outside of New York City (NYC).

**Design:** Interrupted time-series analyses using generalized estimating equations to examine longitudinal school immunization compliance data from the 2012-13 through 2021-22 school years.

**Setting:** New York State public and nonpublic schools outside of NYC.

**Participants:** Schools that submitted ≥1 compliance report in the time periods before and after the legislative repeal of nonmedical exemptions with publicly available student enrollment data.

**Exposure:** NYS Senate Bill 2994A was passed in June 2019, eliminating school entry nonmedical vaccine exemptions; since compliance with the law was evaluated for most students during the following school year, we considered the 2019-20 school year as the law’s effective date.

**Main Outcomes and Measures:** Main outcomes examined were school required vaccine coverage, defined as the percentage of students at each school who completed all grade-appropriate NYS vaccine requirements, and the percentage of students with a medical vaccine exemption.

**Results:** Among 3,525 eligible schools, the implementation of NYS Senate Bill 2994A was associated with an increase in mean required vaccine coverage of 5% and 1% among nonpublic and public schools, respectively, with additional annual increases in coverage observed through the 2021-22 school year. The law’s implementation was also associated with a 0.1% (95% CI: 0.0%, 0.1%) decrease in medical vaccine exemptions at both public and nonpublic schools, and small, but significant mean annual declines in medical vaccine exemptions through the end of the study period.

**Conclusion and Relevance:** The NYS elimination of school entry nonmedical vaccine exemption options was effective to improve required vaccine coverage; coverage gains were not replaced by increases in medical vaccine exemptions.

**KEY POINTS:** *Question:* Was the New York State (NYS) law eliminating nonmedical vaccine exemption options from school entry vaccine requirements effective to increase vaccine coverage among NYS schools (outside of New York City)?

*Findings:* Using interrupted time-series analyses, we found the implementation of the NYS law was associated with an increase in mean required vaccine coverage at NYS schools; small, but significant declines in medical exemptions were also observed in relation to the law.

*Meaning:* State legislation eliminating nonmedical vaccine exemption options from school entry vaccine laws can be effective to improve school vaccine coverage without replacement by medical vaccine exemptions.

## Introduction

Following two large measles outbreaks in New York State (NYS) that threatened the United States’ (US) measles elimination status, the NYS legislature passed Senate Bill 2994A (SB2994A) in June 2019, repealing nonmedical vaccine exemption options from its school entry vaccine requirements.^1–3^ With the law’s passage, New York became the fifth US state to address increasing pediatric undervaccination by eliminating all school nonmedical vaccine exemptions, and the third US state, behind California and Maine, to adopt such a law in recent decades.^4–7^ Since the passage of NYS SB2994A, other states have considered similar legislation, including Connecticut, where nonmedical vaccine exemption options were repealed from school entry vaccine requirements in 2021.^8^ Yet, unlike recent laws passed in California, Maine, and Connecticut, NYS SB2994A became effective immediately, and did not include a grandfather clause provision excusing students with existing nonmedical exemptions from compliance.^8–10^

While school entry vaccine requirements are effective to promote and maintain high US pediatric vaccine coverage,^11, 12^ less is known regarding the effects of legislative repeals of nonmedical vaccine exemption options from these requirements. Evaluations of the California repeal of school nonmedical vaccine exemptions suggest that although the legislation was net effective to increase the total percentage of kindergartners up-to-date for vaccinations, these increases were partially offset by simultaneous increases in student medical vaccine exemptions.^4, 10, 13, 14^ In addition, new medical vaccine exemptions were found to spatially cluster, suggesting that the California legislation did not have a uniform impact on school vaccine coverage.^4, 15^

To date, the effects of other state repeals of nonmedical vaccine exemptions have not yet been evaluated, including in NYS. Further, given differences in the structure, implementation timelines, and target populations of these laws, it is feasible that their impact may differ by jurisdiction. In this study, we evaluated the effects of the NYS SB2994A repeal of nonmedical vaccine exemptions on vaccine coverage and the uptake of medical vaccine exemptions among NYS schools outside of New York City (NYC).

## Methods

### Study setting

NYS SB2994A was signed into law on June 13, 2019.^2^ Although the bill became effective immediately, students were given a 14-day grace period to demonstrate compliance or a pathway towards compliance (*i.e.*, receipt of the first vaccination for all required vaccinations) prior to exclusion from NYS schools.^2^ Due to the timing of this grace period and school summer recess, compliance with the legislation for most students was not assessed until the 2019-20 school year.

### Study design, data sources, and population

We conducted interrupted time-series analyses to examine the effects of NYS SB2994A on annual school required vaccination coverage during the 2012-13 through 2021-22 school years, using the 2019-20 school year as the legislation’s effective date. Annual school vaccination coverage was ascertained using publicly available datasets from the NYS Department of Health (NYSDOH) School Immunization Survey.^16^ Briefly, this annual survey requires schools to report the number of students with medical and religious vaccine exemptions, and those who met grade-appropriate proof of immunity requirements for 7 mandatory school entry immunizations (diphtheria, hepatitis B, measles, mumps, polio, rubella, varicella).^16, 17^ NYSDOH publicly releases aggregated, annual survey data for each school reporting the percentages of students for each variable along with school identifying information. To create a longitudinal dataset, we linked schools across annual survey files using combinations of school ID, name, and address; data were then linked with publicly available school enrollment datasets from the NYS Education Department (NYSED) to identify annual school populations.^18^

Due to differences in governance and reporting structures among New York City schools compared with other NYS locations, NYC schools were excluded from all analyses. Among non-NYC schools, schools must have submitted ≥1 NYSDOH School Immunization Survey in the time periods before (*i.e.*, 2012-2013 through 2018-19 school years) and after (*i.e.*, 2019-20 through 2021-22 school years) the implementation of NYS SB2994A with linkages to annual NYSED enrollment data to be eligible for study inclusion.

### Statistical analyses

In separate analyses, we investigated the effects of NYS SB2994A on the annual percentage of students who completed grade-appropriate requirements for all 7 required vaccines (*i.e.*, required vaccine coverage) and the percentage of students who received a medical exemption at each school as our primary outcomes of interest. In crude analyses, we explored these outcomes as both: (i) unweighted estimates, where each school contributed equally to the analysis, to inform vaccination levels across school clusters, and (ii) weighted estimates, where outcomes were weighted by a school’s student enrollment, to inform vaccination levels across student populations. In adjusted analyses that accounted for the annual repeated sampling of schools, we used binomial generalized estimating equations (GEE) with an identity link function and first-order autoregressive structure to estimate the absolute effect of the law on our primary outcomes with 95% confidence intervals (CIs). In these analyses, the law was modeled as a binary variable, representing 0 and 1 in the time periods before and after the law’s implementation, respectively. Because we anticipated that the law’s effects may change over time, we considered adjustment for a change in trend in the period after the law’s implementation using a product term between the binary law variable and a linear variable accounting for the school year after the law. Additionally, since we anticipated that the law’s effects may differ by school type,^19, 20^ we considered inclusion of a binary variable accounting for nonpublic and public school status and separate product terms to allow for differences in trend by school type in both the time periods before and after the law’s implementation. In secondary analyses, we examined the percentage of students with any vaccine exemptions (*i.e*., nonmedical and medical exemptions) as our outcome variable of interest.

All analyses were conducted using RStudio with R version 4.2.2 (The R Foundation for Statistical Computing, Vienna, Austria).

## Results

### Study population

Figure 1 depicts a flow diagram of annual NYSDOH School Immunization Survey reports considered for study inclusion. Briefly, we identified 3,938 distinct non-NYC, NYS schools (2,907 public, 1,031 nonpublic) that completed ≥1 survey during the study period. Of these, 3,525 (89.5%) schools submitted ≥1 survey during the time periods before and after the law’s implementation, with linkages to NYSED enrollment data. Among these eligible schools, 2,733 (94%) public and 792 (77%) nonpublic schools were represented, submitting a mean of 9.3 (95% CI: 9.3, 9.4) and 7.9 (95% CI: 7.8, 8.1) reports across the 10-year study period, respectively. Stratified by time period, public and nonpublic schools submitted a mean of 6.4 (95% CI: 6.4, 6.5) and 5.4 (95% CI: 5.3, 5.6) reports, respectively, during the 7 school years prior to SB2994A implementation; during the 3-year period following the law, these schools submitted a mean of 2.9 (95% CI: 2.9, 2.9) and 2.6 (95% CI: 2.5, 2.6) reports, respectively. A total of 31,718 distinct surveys from all schools were included in analyses.

**Figure 1.**
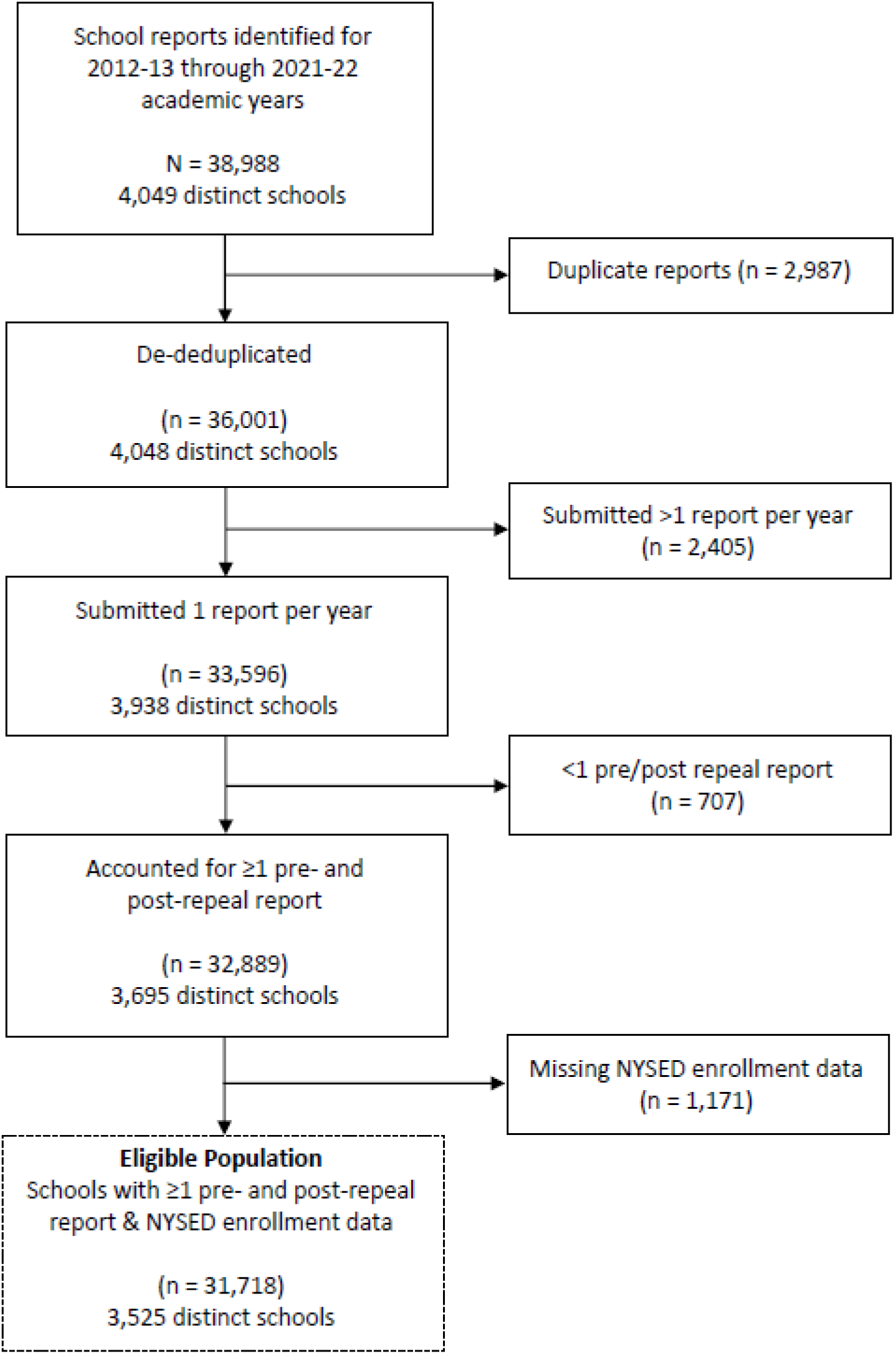
Flow diagram of included schools. *Legend:* NYSED = New York State Education Department.

### Completion of required vaccines

Table 1 presents crude analyses of mean required vaccine coverage. At the beginning of the study period (2012-13 school year), overall mean coverage weighted by student enrollment was 98.4% (95% CI: 98.2%, 98.5%) and fell to 97.0% (95% CI: 96.7%, 97.2%) in 2018-19, or the school year prior to NYS SB2994A implementation; during this same time period, unweighted coverage started at 96.8% (95% CI: 96.4%, 97.1%) and fell to 95.1% (95% CI: 94.7%, 95.5%). In both weighted and unweighted estimates, vaccine uptake was consistently lower among nonpublic schools during the pre-implementation period. In the year before NYS SB2994A implementation (2018-19), weighted coverage was 97.6% (95% CI: 97.5%, 97.7%) and 90.8% (95% CI: 88.6%, 93.0%) among public and nonpublic school students, respectively, while unweighted coverage was 97.6% (95% CI: 97.5%, 97.7%) and 85.7% (95% CI: 84.1%, 87.2%) at public and nonpublic schools, respectively. Following the law’s implementation, weighted mean vaccine coverage increased by 0.7% (95% CI: 0.5%, 0.9%) and 4.1% (95% CI: 1.8%, 6.5%) among public and nonpublic school students, respectively, while unweighted coverage increased by 0.6% (95% CI: 0.4%, 0.8%) at public schools and 3.7% (95% CI: 1.5%, 5.9%) at nonpublic schools. Small, annual gains in vaccine coverage were also observed through the end of the study period (2021-22), with weighted mean vaccine coverage of 98.4% (95% CI: 98.3%, 98.5%) and 96.3% (95% CI: 95.5%, 97.1%) among public and nonpublic school students, and unweighted estimates of 98.5% (95% CI: 98.4%, 98.6%) and 92.4% (95% CI: 91.0%, 93.9%) among public and nonpublic schools.

**Table 1.** Required vaccine coverage among New York State (NYS) schools (excluding New York City) before and after the NYS Senate Bill (SB) 2994A repeal of nonmedical exemptions.

| Start of Academic Year |  | Mean % Required Vaccinations: Unweighted Percentages |  |  | Mean % Required Vaccinations: Weighted Percentages |  |  |
| --- | --- | --- | --- | --- | --- | --- | --- |
|  |  | Overall | Public | Nonpublic | Overall | Public | Nonpublic |
| Pre-SB 2994A | 2012 | 96.8% | 98.5% | 88.9% | 98.4% | 98.6% | 95.1% |
|  | 2013 | 96.7% | 98.5% | 88.9% | 98.3% | 98.6% | 94.9% |
|  | 2014 | 95.9% | 97.7% | 88.2% | 97.6% | 97.9% | 94.1% |
|  | 2015 | 95.8% | 97.8% | 87.8% | 97.6% | 98.0% | 93.7% |
|  | 2016 | 95.5% | 97.6% | 87.2% | 97.0% | 97.5% | 92.6% |
|  | 2017 | 95.0% | 97.3% | 86.1% | 96.8% | 97.3% | 91.4% |
|  | 2018 | 95.1% | 97.6% | 85.7% | 97.0% | 97.6% | 90.8% |
| Post-SB 2994A | 2019 | 96.5% | 98.2% | 89.4% | 98.0% | 98.3% | 94.9% |
|  | 2020 | 97.7% | 99.0% | 93.2% | 98.7% | 99.0% | 96.1% |
|  | 2021 | 97.4% | 98.5% | 92.4% | 98.2% | 98.4% | 96.3% |

Final selected GEE models examining school required vaccine coverage contained terms for school type and separate product terms to allow for differences in trend by school type in both the pre- and post-implementation periods (Supplemental Appendix 1). Figure 2 displays modelled vaccine coverage estimates. In general, trends were similar to crude analyses. Briefly, baseline coverage was higher among public versus nonpublic schools; while both school types experienced annual declines in coverage during the pre-implementation period, declines in mean coverage among nonpublic schools were greater. Upon implementation of SB2994A, mean required vaccine coverage increased by 0.8% (95% CI: 0.7%, 1.0%) among public schools and 4.6% (95% CI: 3.7%, 5.6%) among nonpublic schools. Both public and nonpublic schools continued to experience annual increases of 0.3% (95% CI: 0.2%, 0.4%) and 1.3% (95% CI: 0.8%, 1.7%), respectively, during the post-implementation period. Supplemental Appendix 1 includes a table of model results.

**Figure 2.**
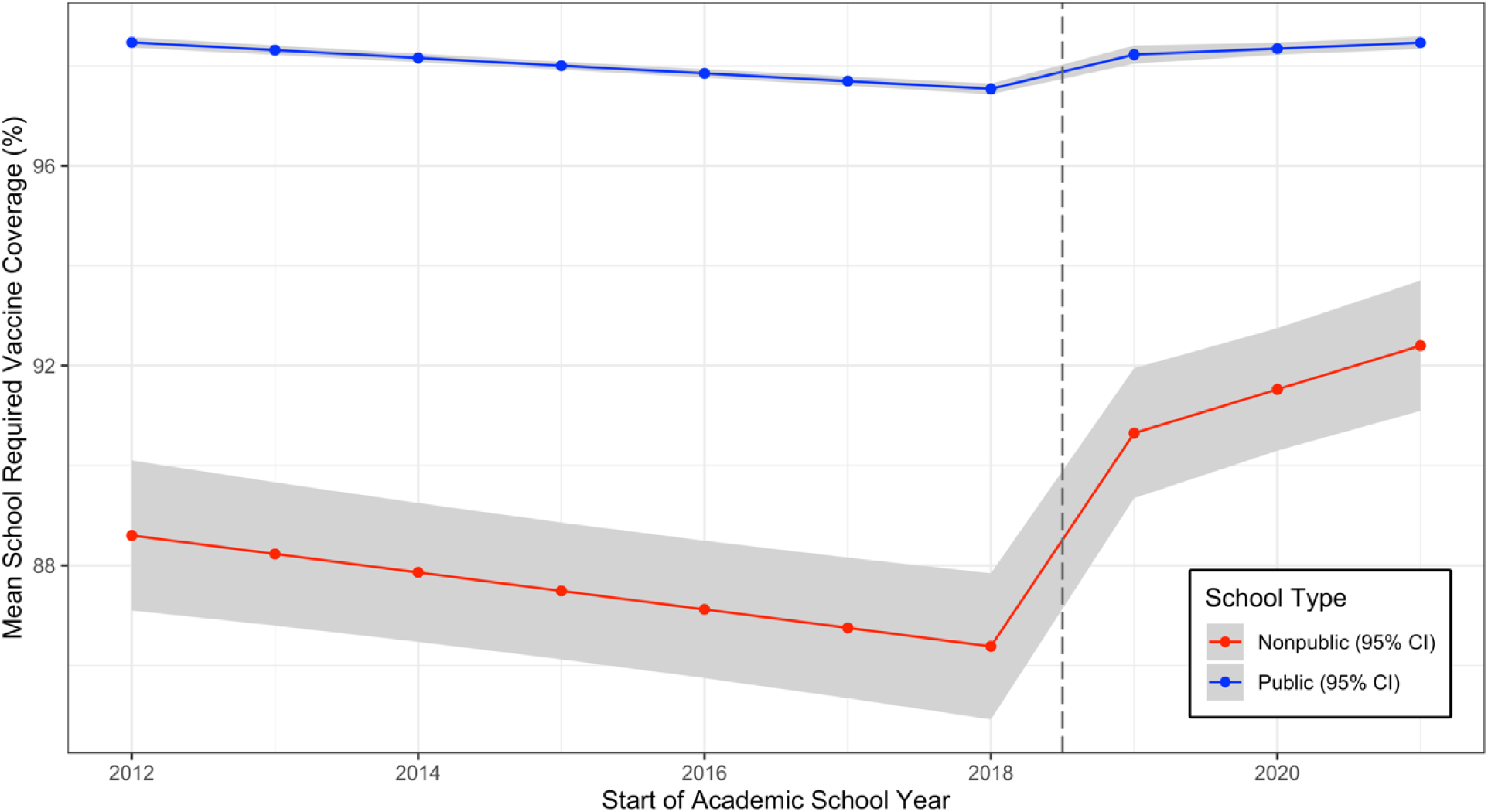
Generalized estimating equations (GEE) modeled estimates of school required vaccine coverage by school type. The New York State Senate Bill 2994A repeal of nonmedical exemptions affected students in the 2019-20 school year. *Legend:* CI = confidence interval.

### Uptake of vaccine exemptions

Figure 3 examines weighted and unweighted mean medical vaccine exemption uptake among public and nonpublic schools over time. In both weighted and unweighted analyses, medical exemptions were more common among nonpublic schools compared to public schools during the pre-implementation period and increased slightly across the time period prior to the law’s implementation. However, in general, medical exemptions were rare, accounting for a weighted average of 0.2% (95% CI: 0.2%, 0.2%) and 0.3% (95% CI: 0.2%, 0.3%) of public and nonpublic school students, respectively, in the year prior to SB2994A implementation. Following the repeal, the weighted average of medical exemptions among public and nonpublic school students remained unchanged. Final GEE models examining this outcome contained a term for school type and separate terms to allow for differences in trend in both the pre- and post-implementation periods (Supplemental Appendix 2). Among public and nonpublic schools, a small, but significant increase in medical vaccine exemptions was observed in the time period before NYS SB2994A. In the school year following the law’s implementation, these schools experienced a 0.1% (95% CI: 0.0%, 0.1%) decrease in medical vaccine exemptions, with annual declines of 0.02% (95% CI: 0.01%, 0.03%) observed through the end of the study period.

**Figure 3.**
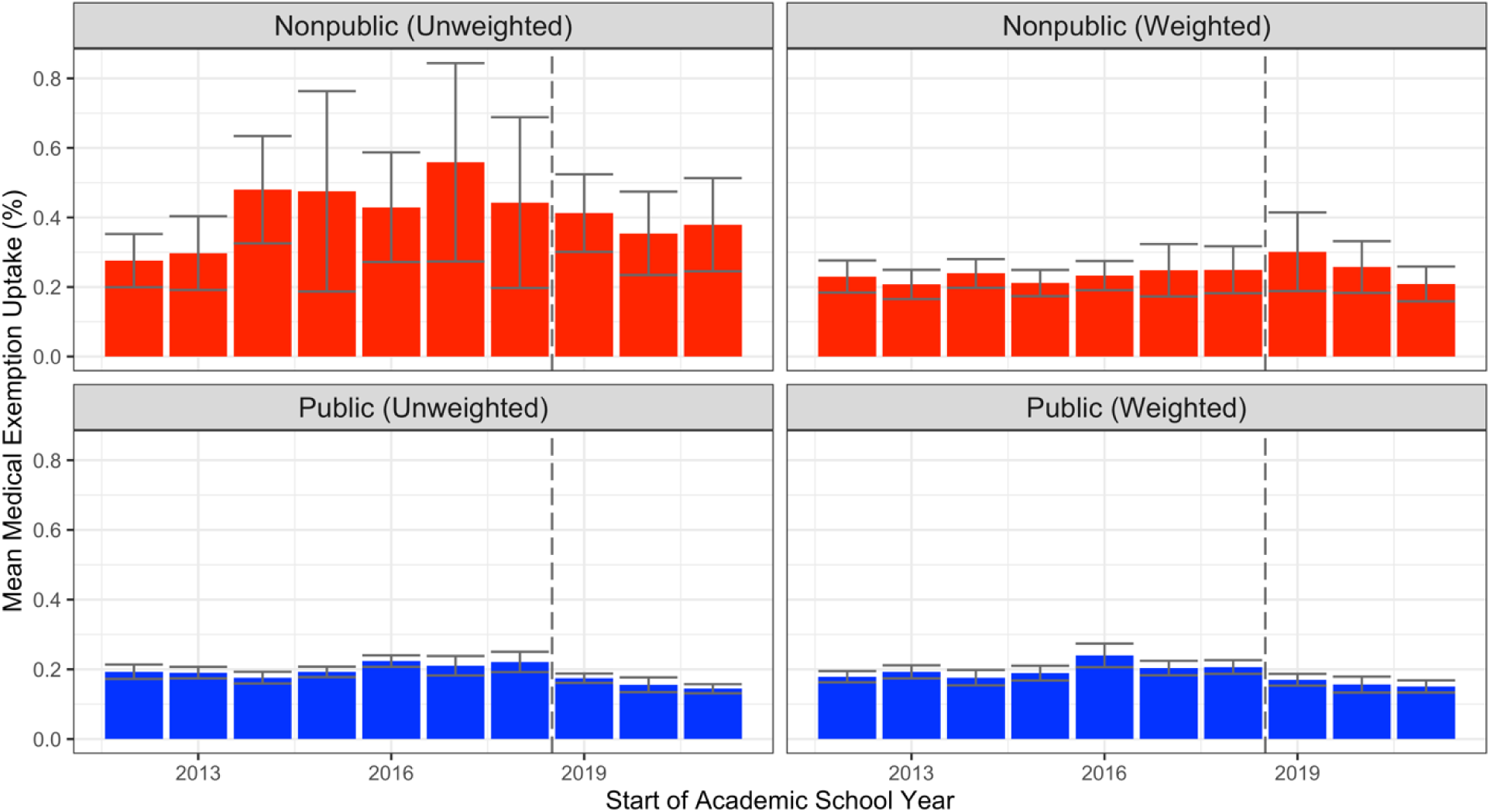
Medical vaccine exemption uptake by school type. The New York State (NYS) Senate Bill (SB) 2994A repeal of nonmedical exemptions affected students in the 2019-20 school year. Variables represent weighted and unweighted mean percentages with 95% confidence intervals (CI).

Figure 4 examines crude data for the uptake of any vaccine exemptions over time by school type (*i.e.,* public or nonpublic). Overall, weighted estimates found 1.0% (95% CI: 1.0%, 1.0%) of public students and 3.4% (95% CI: 2.9%, 4.0%) of nonpublic students reported a vaccine exemption in the year prior to NYS SB2994A implementation; unweighted estimates found mean uptake of vaccine exemptions was 1.1% (95% CI: 1.0%, 1.1%) among public schools and 8.4% (95% CI: 7.1%, 9.7%) among nonpublic schools. Following NYS SB2994A implementation, the mean percentage of school vaccine exemptions decreased in both weighted and unweighted estimates. At the end of the study period (2021-22), the weighted uptake of any vaccine exemptions was 0.2% (95% CI: 0.1%, 0.2%) among public school students and 0.2% (95% CI: 0.2%, 0.3%) among nonpublic school students, with unweighted estimates of 0.1% (95% CI: 0.1%, 0.2%) and 0.4% (95% CI: 0.2%, 0.5%) among public and nonpublic schools, respectively. Final GEE models examining this outcome contained terms for school type and separate product terms to allow for differences in trend by school type in both the pre- and post-implementation periods (Supplemental Appendix 3), with similar results in comparison with crude analyses. Upon the law’s implementation, public schools experienced a 1.0% (95% CI: 1.0%, 1.0%) decline in all vaccine exemptions, while nonpublic schools experienced an 8.1% (95% CI: 6.8%, 9.4%) decline; additional annual declines of 0.1% (95% CI: 0.1%, 0.1%) and 0.3% (95% CI: 0.2%, 0.4%) among these schools, respectively, were observed through the 2021-22 school year.

**Figure 4.**
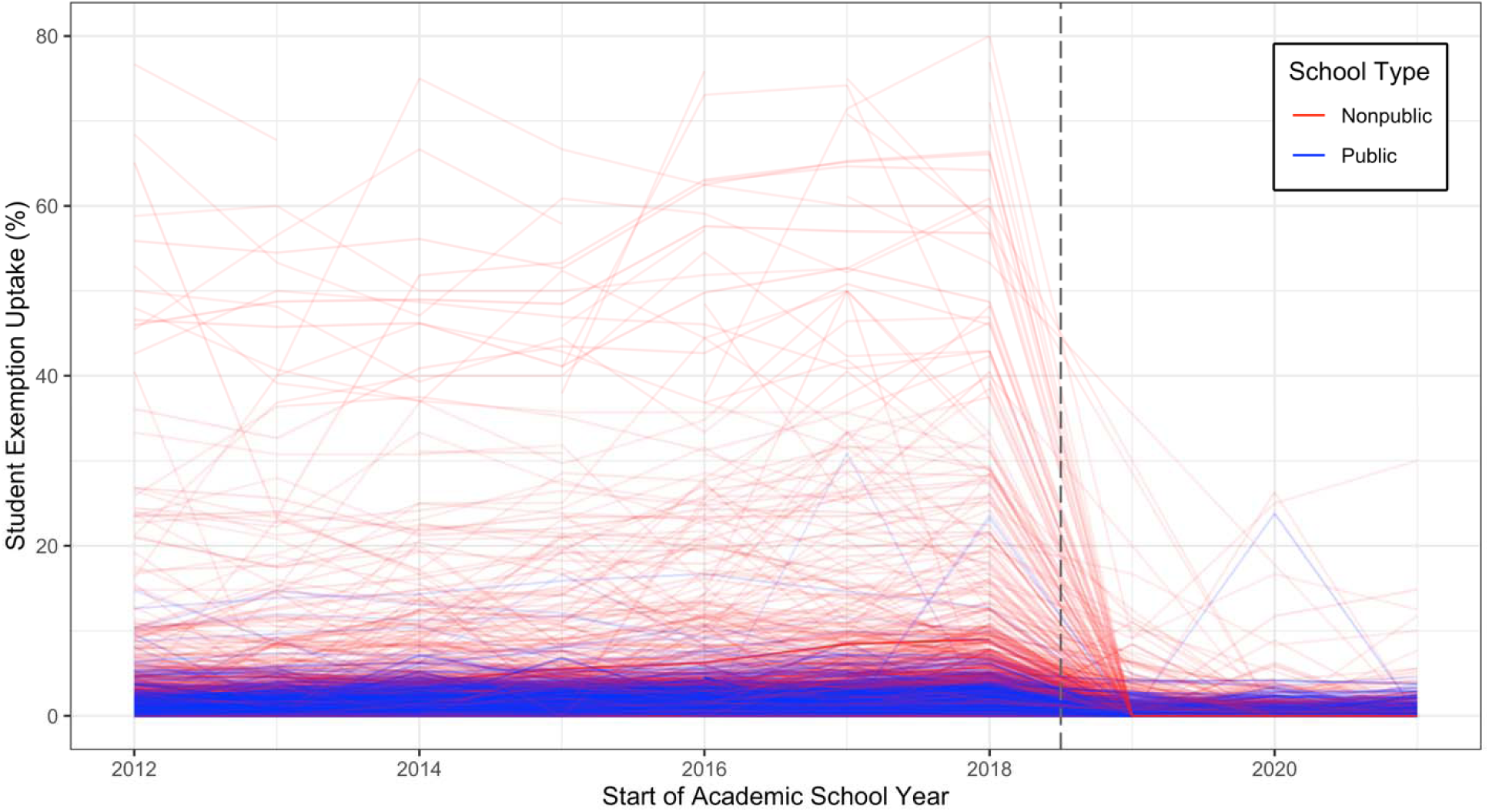
Uptake of school vaccine exemptions by school type. Line transparency has been adjusted by student enrollment, with lower enrollment depicted as a more transparent line. For visualization purposes, vaccine exemption percentages above 80% are not displayed.

## Discussion

Using interrupted time-series analyses, we examined the effects of NYS SB2994A on required vaccine coverage and the uptake of medical vaccine exemptions among non-NYC, NYS schools. Prior to NYS SB2994A implementation, we observed consistent declines in school required vaccine coverage over time, with unweighted coverage of 85.7% among nonpublic schools and 97.6% among public schools in the school year prior to the law’s adoption. Using longitudinal GEE models, we estimated that NYS SB2994A implementation was associated with absolute increases in mean required vaccine coverage of 4.6% among nonpublic schools and 0.8% among public schools, with additional, annual mean increases of 1.2% among nonpublic schools and 0.3% among public schools estimated through the 2021-22 school year. Simultaneously, we also found that NYS SB2994A implementation was associated with a small, but significant absolute decline of 0.1% in medical vaccine exemption uptake among nonpublic and public schools, with subsequent annual mean declines of 0.02% among these schools through the 2021-22 school year. Collectively, these results demonstrate that the NYS SB2994A repeal of nonmedical vaccine exemptions was effective to address declining required vaccine coverage at non-NYC, NYS schools, and that coverage gains were not replaced by increases in the uptake of medical vaccine exemptions.

To our knowledge, this is the first study to evaluate the impact of NYS SB2994A on required vaccine coverage and the uptake of medical exemptions. While evaluations of the repeal of nonmedical exemptions in California suggest that declines in school nonmedical vaccine exemptions were partially offset by the uptake of new medical vaccine exemptions,^4, 10, 13, 14^ we found that non-NYC, NYS schools did not share these experiences. Since NYS SB2994A did not change school policies regarding medical exemptions, we hypothesize that the small, but significant declines we observed in medical vaccine exemptions after the implementation of the law may be due to the adoption of stricter documentation or validation requirements for these exemptions by schools. Conversely, our results may also suggest that increasing medical exemptions observed after the California repeal of nonmedical vaccine exemptions may not solely be related to the repeal of nonmedical exemptions, but rather, could be an artifact of the structure of the California legislation. For example, the California Senate Bill 277 (SB277) that repealed nonmedical vaccine exemptions contained explicit provisions broadening the use of medical exemptions for reasons that do not represent contraindications to vaccination, such as a family medical history.^5, 14^ The apparent differences between our findings related to medical exemptions and the experiences of California may have important implications for policymakers in other jurisdictions considering similar legislation. Nonetheless, additional research to examine longitudinal trends and spatial clustering of medical exemptions among NYS schools is warranted to understand whether these findings remain stable into the future.

Our crude analyses examining weighted versus unweighted mean required vaccine coverage demonstrate the public health challenges of addressing undervaccination to prevent vaccine-preventable disease outbreaks in the population. While our weighted estimates demonstrate high overall required vaccine coverage of 97% among NYS school children in the year prior to NYS SB2994A implementation, unweighted mean required vaccine coverage during the same time period was only 86% at nonpublic schools. This contrast highlights apparent clustering of vaccine exemptions by school, which has important implications for the control and spread of vaccine-preventable diseases in the population. Furthermore, although our study provides strong evidence that NYS SB2994A was effective to increase mean vaccine coverage, we also found differences in required vaccine coverage between public and nonpublic schools persisted, with public schools having 2% higher mean weighted required vaccine coverage or 6% higher mean unweighted required vaccine coverage in comparison with nonpublic schools at the end of our study period. While these differences by school type were narrowed in comparison with the year prior to NYS SB2994A implementation, these gaps may suggest greater susceptibility of nonpublic schools to vaccine-preventable disease outbreaks.

## Limitations

Our results should be considered with regard to several limitations. First, our study period is limited to 3 school years following NYS SB2994A implementation, which includes years affected by the COVID-19 pandemic. Due to these limitations, our results may not represent longitudinal school vaccination trends, and NYS SB2994A effects should be evaluated into the future. Nonetheless, these limitations are partially offset by our study design, which allowed us to consider longitudinal trends in school vaccine coverage in the 7 years prior to the implementation of the law; further, based on the timing of the COVID-19 pandemic and school closures in March 2020, initial vaccine coverage for the 2019-20 school year, or the first year following NYS SB2994A implementation, was less likely to be affected by the pandemic. Second, due to differences in reporting and governance structures, our analyses did not include NYC schools; therefore, our findings may not be generalizable to this region. Third, 10% of non-NYC schools that submitted ≥1 compliance report during the 10-year study period were excluded from our analyses because they did not meet study eligibility criteria.

Should these schools have differing experiences with the legislation, our findings may be biased; however, our analyses remain important as they represent the experiences of 90% of non-NYC, NYS schools. Finally, our analyses did not examine whether NYS SB2994A was associated with changes in school enrollment. Where NYS SB2994A prompted unvaccinated students to leave schools to avoid compliance, these changes could have implications for the spread of vaccine-preventable diseases in the population; future research is necessary to examine these potential effects.

## Conclusions

Using interrupted time-series analyses, we found that the NYS SB2994A repeal of nonmedical exemptions was associated with increases in mean required vaccine coverage at non-NYC NYS schools. Among these schools, we did not find evidence that coverage gains were offset by increasing medical vaccine exemptions; instead, we observed a small, but significant decline in medical vaccine exemptions associated with the law. These findings suggest that the legislative repeal of school entry nonmedical vaccine exemptions can be effective to improve vaccination compliance without replacement by new medical vaccine exemptions. Although our study period includes 3 years of data following the implementation of NYS SB2994A, continued research is necessary to examine the law’s long-term effects.

## Data Availability

All data used in these analyses are publicly available from the New York State Department of Health and the New York State Education Department.

