## Supplemental Appendix for "Evaluation of School Vaccine Coverage and Medical Vaccine Exemptions Following the Repeal of School Entry Nonmedical Vaccine Exemption Options in New York State"

**Supplemental Appendix 1.** Final selected binomial generalized estimating equations (GEE) model with an identity link function and results estimating the impact of New York State (NYS) Senate Bill 2994A on required immunization completion.

| **Final selected model:** | | | | |
| --- | --- | --- | --- | --- |
| $Complete {vx}_{ij}= \beta_{0}+\beta_{1}{Time}_{ij}+\beta_{2}School {type}_{i}+\beta_{3}{Time}_{ij}*School {type}_{i}+\beta_{4}{Law}_{ij}+\beta_{5}{Law}_{ij}*School{type}_{i}+\beta_{6}Time post {law}_{ij}+\beta_{7}Time post {law}_{ij}*School{type}_{i}+\epsilon_{ij}$ | | | | |
| **Term** | **Estimate** | **P value** | **Lower CI** | **Upper CI** |
| Intercept | 0.9847 | <0.001 | 0.9837 | 0.9858 |
| Time | -0.0016 | <0.001 | -0.0018 | -0.0013 |
| Law | 0.0084 | <0.001 | 0.0067 | 0.0101 |
| Time post law | 0.0027 | <0.001 | 0.0018 | 0.0037 |
| School type | -0.0987 | <0.001 | -0.0840 | -0.1135 |
| Time$*$School type | -0.0021 | 0.0245 | -0.0003 | -0.0040 |
| Law$*$School type | 0.0380 | <0.001 | 0.0282 | 0.0477 |
| Time post law$*$School type | 0.0097 | <0.001 | 0.0048 | 0.0146 |
| **Where:**   - *Complete vx_ij_ =* Mean school required vaccine completion (%) - *Time_ij_* = Time from the start of study period (years) - *Law_ij_ =* NYS Senate Bill 2994A implementation (0=pre-implementation; 1=post-implementation) - *Time post law_ij_* = Time from Senate Bill 2994A implementation (school years) - *School type_ij_* = Represents public or nonpublic school (0=public or 1=nonpublic) - $\epsilon$ = Error term - *i* = Individual school - *j* = Time | | | | |

**Supplemental Appendix 2.** Final selected binomial generalized estimating equations (GEE) model with an identity link function and results estimating the impact of New York State (NYS) Senate Bill 2994A on medical exemption uptake.

| $Medical exemption {uptake}_{ij}= \beta_{0}+\beta_{1}{Time}_{ij}+\beta_{2}School {type}_{i}+\beta_{4}{Law}_{ij}+\beta_{6}Time post {law}_{ij}+\epsilon_{ij}$ | | | | |
| --- | --- | --- | --- | --- |
| **Term** | **Estimate** | **P value** | **Lower CI** | **Upper CI** |
| Intercept | 0.0018 | <0.001 | 0.0016 | 0.0020 |
| Time | 0.0001 | 0.0296 | 0.0000 | 0.0001 |
| Law | -0.0006 | 0.0053 | -0.0002 | -0.0010 |
| Time post law | -0.0002 | <0.001 | -0.0001 | -0.0003 |
| School type | 0.0020 | <0.001. | 0.0011 | 0.0029 |
| **Where:**   - *Medical exemption uptake_ij_* = Mean school medical exemption uptake (%) - *Time_ij_* = Time from the start of study period (years) - *Law_ij_ =* NYS Senate Bill 2994A implementation (0=pre-implementation; 1=post-implementation) - *Time post law_ij_* = Time from Senate Bill 2994A implementation (years) - *School type_ij_* = Represents public or nonpublic school (0=public or 1=nonpublic) - $\epsilon$ = Error term - *i* = Individual school - *j* = Time | | | | |

**Supplemental Appendix 3.** Final selected binomial generalized estimating equations (GEE) model with an identity link function and results estimating the impact of New York State (NYS) Senate Bill 2994A on the uptake of all vaccine exemptions.

| $Overall exemption {uptake}_{ij}= \beta_{0}+\beta_{1}{Time}_{ij}+\beta_{2}School {type}_{i}+\beta_{3}{Time}_{ij}*School {type}_{i}+\beta_{4}{Law}_{ij}+\beta_{5}{Law}_{ij}*School{type}_{i}+\beta_{6}Time post {law}_{ij}+\beta_{7}Time post {law}_{ij}*School{type}_{i}+\epsilon_{ij}$ | | | | |
| --- | --- | --- | --- | --- |
| **Term** | **Estimate** | **P value** | **Lower CI** | **Upper CI** |
| Intercept | 0.0067 | <0.001 | 0.0063 | 0.0070 |
| Time | 0.0007 | <0.001 | 0.0006 | 0.0008 |
| Law | -0.0098 | <0.001 | -0.0093 | -0.0103 |
| Time post law | -0.0008 | <0.001 | -0.0007 | -0.0009 |
| School type | 0.0525 | <0.001 | 0.0407 | 0.0644 |
| Time$*$School type | 0.0028 | <0.001 | 0.0018 | -0.0039 |
| Law$*$School type | -0.0712 | <0.001 | -0.0582 | -0.0839 |
| Timepost law$*$School type | --0.0024 | <0.001 | -0.0012 | -0.0036 |
| **Where:**   - *Overall exemption uptake_ij_ =* Mean school medical and religious exemption uptake (%) - *Time_ij_* = Time from the start of study period (years) - *Law_ij_ =* Represents period before and after Senate Bill 2994A implementation (0 or 1) - *Time post law_ij_* = Time from Senate Bill 2994A implementation (years) - *School type_ij_* = Represents public or nonpublic school (0 or 1) - $\epsilon$ = Error term - *i* = Individual school - *j* = Time | | | | |
